# From iRBD to Parkinson’s disease: Tracking Glymphatic Dysfunction Using Automated DTI-ALPS Analysis

**DOI:** 10.1101/2025.01.26.25321139

**Authors:** S Marecek, V Rottova, J Nepozitek, T Krajca, R Krupicka, J Keller, D Zogala, J Trnka, K Sonka, E Ruzicka, P Dusek

## Abstract

Diffusion tensor image analysis along the perivascular space (DTI-ALPS) is a non-invasive marker of glymphatic function that typically relies on manual region of interest (ROI) placement. This study compared glymphatic function in treatment-naïve, de novo diagnosed patients with Parkinson’s disease (PD), patients with isolated REM behavior disorder (iRBD), and healthy controls using both manual and automatic DTI-ALPS methods. ALPS scores were analyzed bilaterally and correlated with clinical severity (MDS-UPDRS) and nigrostriatal denervation (DAT-SPECT). The study included 79 PD patients (60±12 years), 57 iRBD patients (67±7 years), and 48 controls (62±10 years). ANCOVA revealed significant inter-group differences using both manual (p=0.018) and automatic (p=0.002) methods. Automatic analysis showed significantly lower ALPS scores in PD compared to controls (p=0.001) and iRBD (p=0.009). ALPS scores correlated with symptom severity and nigrostriatal degeneration. These findings highlight early glymphatic dysfunction in PD and demonstrate the reliability of the automatic DTI-ALPS method.

## Introduction

The glymphatic system is a whole-brain perivascular network that utilizes periarterial cerebrospinal fluid (CSF) influx, with its subsequent diffusion into the brain parenchyma and perivenous efflux to drive interstitial solute clearance.^1,2^ Dysfunction of the glymphatic system is increasingly recognized as a contributing factor in several neurological disorders, including Parkinson’s disease (PD), normal pressure hydrocephalus, Alzheimer’s disease, and others^3–6^. The pathophysiological mechanisms of glymphatic dysfunction in PD are not well known. However, one study in mice has shown α- synuclein injection into basal ganglia caused delayed dural lymphatic vessel drainage, and vice versa, the ligation of draining lymphatic vessels caused increased α-synuclein accumulation and exacerbated motor and memory deficits^7^. In PD, glymphatic dysfunction has been observed even in the earliest stages of neurodegeneration, including its prodromal stages, represented by isolated rapid-eye-movement behavior disorder (iRBD)^8,9^.

RBD is a disorder characterized by dream enactment and the loss of muscle atonia during REM sleep. It can be caused by narcolepsy, antidepressant intake, brainstem lesions, and, in cases when a precise reason has not been found, RBD is termed “isolated”. This isolated RBD (iRBD) is predominantly caused by early stage synucleinopathies, with studies showing up to 90% conversion rate in 14 years to either PD, dementia with Lewy bodies, or multiple system atrophy. This makes iRBD a valuable target for studying PD’s pathophysiology, course, and possible therapy^10,11^.

There are several ways to measure the function of the glymphatic system^12^, with diffusion tensor imaging along perivascular spaces (DTI-ALPS) being one of them, first described in 2017 by Taoka et. al^13^. This method uses widely available MRI sequences and relies on the placement of regions of interest (ROI) next to the top of the lateral ventricles, into the areas of projection and association fibers. In this area the glymphatic flow traverses the perivascular spaces, perpendicular to the lateral ventricles and both the projection and association fibers, thus presenting a unique opportunity to measure the flow by the means of diffusion tensor imaging^6^.

According to our best knowledge, only two studies simultaneously compared patients with PD, iRBD, and healthy controls^9,14^. In one of these studies, iRBD patients were diagnosed based on history rather than polysomnography, classifying them as “possible” iRBD cases^14^. In the other study, each group was limited to only 20 participants^9^. In our study, we aim to compare the glymphatic function as measured by DTI-ALPS scores in newly diagnosed treatment naïve patients with PD, patients with video-polysomnography confirmed iRBD and healthy controls. Additionally, we explore the relationship between the ALPS index, clinical scores and nigrostriatal dopaminergic degeneration measured by dopamine transporter single-photon emission computed tomography (DAT-SPECT).

Furthermore, most of the studies using the DTI-ALPS method employ a manual selection of regions of interest (ROIs)^6^. This introduces a possibility of human error or bias. We aimed to employ an automatic ROI selection algorithm that would forego the need for selection of these ROIs by clinicians.

## Methods

### Participants

Our subject population consisted of de novo diagnosed treatment-naïve subjects with PD, iRBD subjects, and healthy controls recruited at the Department of Neurology, First Faculty of Medicine, Charles University and General University Hospital in Prague during 2015-2021. The PD patients were part of the BIO-PD cohort described previously^15^ and were diagnosed according to the Movement Disorders Society (MDS) clinical diagnostic criteria^16^. The iRBD patients were diagnosed in accordance with the International Classification of Sleep Disorders, third edition (ICSD-3) using video-polysomnography^17^. Patients with RBD secondary to focal brainstem lesions, narcolepsy, medication usage, and with clinically manifest dementia or parkinsonism, were excluded. The control subjects were recruited from the general community via advertisements. Eligibility criteria included the absence of active oncologic illness, significant neurological disorders, and abuse of psychoactive substances. RBD was excluded in all control subjects by history and video-polysomnography. All study participants were examined according to a complex protocol including neurological examination, structured interview, Montreal Cognitive Assessment (MoCA)^18^, and MDS-sponsored Revision of the Unified Parkinson’s Disease Rating Scale (MDS-UPDRS)^19^. The study was approved by the local Ethics Committee and participants signed informed consent before entering the study, in accordance with the Helsinki Declaration.

### Imaging acquisition protocol

MRI examination was performed on a 3T scanner (Siemens Skyra 3T, Siemens Healthcare, Erlangen, Germany) with a 32-channel head coil. The protocol included diffusion tensor MRI with repetition time (TR) = 10.5 s; echo time (TE) = 93 ms; total 72 slices with isotropic voxel resolution of 2 mm^3^; 30 noncolinear directions with b-value of 1000 s/m^2^ and one b=0 s/m^2^ image in the anterior-posterior and posterior-anterior phase encoding directions; and an axial 3D T1-weighted Magnetization Prepared Rapid Gradient Echo (MPRAGE, TR 2,200 ms; TE 2.4 ms; inversion time (TI) 900 ms; flip angle (FA) 8°; field of view (FOV) 230×197×176 mm; isotropic voxel resolution 1 mm^3^).

In all PD patients and all but four iRBD patients, DAT-SPECT was performed using the [123I]-2-b- carbomethoxy-3b-(4-iodophenyl)-N-(3-fluoropropyl) nortropane ([123I]FP-CIT, DaTscan®, GE Healthcare, Little Chalfont, Buckinghamshire, UK) tracer according to European Association of Nuclear Medicine (EANM) procedure guidelines^20^; the detailed protocol is described elsewhere^21^. Automated semi-quantitative analysis was performed using the DaTQUANT v. 2.0 software (GE Healthcare, USA), and the Z-scores of specific binding ratios (SBR) in both putamina relative to background binding were calculated. DAT-SPECT was performed within one month from MRI. All PD patients were scanned before the introduction of dopaminergic therapy.

### Calculating the DTI-ALPS index

For general image preprocessing we used FSL^22,23^ and MRTrix3^24,25^ software. We have created an automatic pipeline in Snakemake software^26,27^, with the most resource-intensive computing being done on the CESNET MetaCentrum distributed computing infrastructure. The whole analysis was done in each subject’s native diffusion space and the DWI data were denoised, Gibbs ringing artifacts were removed, images were corrected for distortion, eddy currents, and movement. The resulting images were further processed using FSL’s DTIFIT tool, acquiring the Dxx, Dyy, Dzz tensor images.

The DTI-ALPS method relies on placing regions of interest (ROIs) in the areas of association and projection fibers next to the top of lateral ventricles. In these regions, glymphatic flow moves along the x-axis, association fibers run along the y-axis and projection fibers descend along the z-axis. The ALPS-index is calculated by averaging diffusivity along the x-axis, where glymphatic flow occurs, and dividing it by the average diffusivity along the axes that do not contain glymphatic flow or white matter fibers - specifically, the y-axis for the projection area and the z-axis for the association area (1).

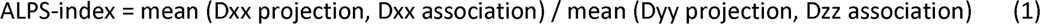

We chose to employ two methods of ROI selection – manual and automatic.

For the classical manual approach, a blinded rater (V.R.) placed 6x6x6 mm^3^ ROIs in the projection and association white matter regions of both hemispheres.

For automatic approach (Figure 1) we used the “JHU ICBM tracts maxprob thr25 1mm” atlas’s labels of superior longitudinal fasciculus (SLF) and corticospinal tract (CST) for the association and projection regions, respectively^28–30^. We restricted these labels to the area on the top of the lateral ventricles and excluded parts close to the cortex, as these could potentially intrude into the cortical area during transformation to the subject’s diffusion space. We then created a single transformation from MNI-152 space to each subject’s diffusion space in a three step process using FSL: 1) we used FLIRT’s linear transformation to register each subject’s b=0 diffusion image to their structural t1 image, 2) we used FLIRT and FNIRT’s non-linear transformation to register patients’ t1 images to the MNI-152 template, and 3) we created a single transformation from the MNI-152 template to each subject’s diffusion space using CONVERTWARP. Subsequently, we transformed the restricted SLF and CST ROIs into each subject’s diffusion space and binarized them, thus generating the association and projection ROI masks subsequently used in the ALPS-index calculation.

**Figure 1.**
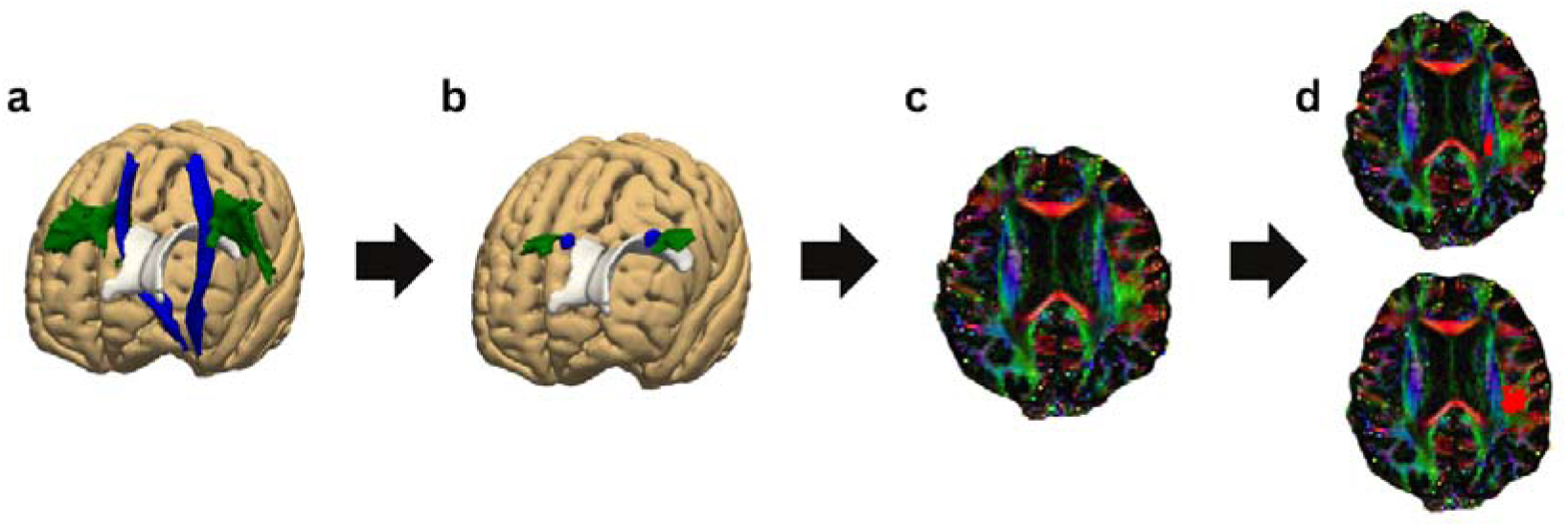
Automatic ALPS ROI selection We used the “JHU ICBM tracts maxprob thr25 1mm” atlas’s labels of superior longitudinal fasciculus and corticospinal tract, for the association and projection areas, respectively (a). We restricted these labels to the area on the top of the lateral ventr icles and excluded parts close to the cortex (b). We then calculated a warp from the MNI152 space to each subject’s diffusion space ( c) and transformed the masks (d). The images presented in steps (c) and (d) display fractional anisotropy-modulated diffusivity in the x (red), y (green), and z (blue) directions.

### Statistics

Statistical analyses were performed using IBM SPSS for Windows, Version 26. We used the Chi- squared test to compare the sex distribution between groups and a one-way ANOVA to explore possible inter-group differences in age. We used one-way ANCOVA with age and sex as covariates to calculate the differences in MDS-UPDRS and MoCA scores. Given the known association of ALPS and age^31^, the ALPS scores were adjusted for age using the regression coefficient β and Age from control subjects as follows^32^:

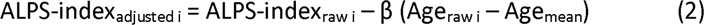

Using one-way ANCOVA with sex as covariate, we determined the inter-group differences in the age- adjusted ALPS scores. All post-hoc multiple comparisons were performed using the least significant difference. The measure of effect size was expressed as partial eta squared (partial η^2^).

We used partial correlation with sex as a covariate to ascertain the correlation between age-adjusted ALPS scores and the mean putaminal SBR Z-scores from both hemispheres; we used age and sex as covariates to determine the correlations with MoCA and MDS-UPDRS scores. We also utilized this method to determine the correlation between ALPS scores and following MDS-UPDRS III subscores: tremor (sum of items 15–18) bradykinesia (sum of items 2, 4-9 and 14), rigidity (item 3), and axial (sum of items 1 and 9-13) subscores^33^.

To assess the possible side differences of ALPS scores based on the magnitude of nigrostriatal denervation, we used the paired Student’s t-test to compare the side with higher and the side with lower putaminal SBR Z-score on DAT-SPECT.

As the iRBD group was significantly older than the PD and control groups, and had significantly more males than the PD group, we performed a sensitivity analysis in age- and sex-matched subgroups to exclude potential bias. The matching was performed using the SPSS’ case control matching utility, with randomized case order when drawing matches and a match tolerance value of 8 for age and 0 for sex. We performed identical age adjustment for ALPS scores as described above.

To compare the manual and automatic approach in all parameters (mean, right-hemisphere and left-hemisphere ALPS scores) we applied three statistical methods: 1) a paired Student’s t-test to assess significant differences, 2) a bivariate correlation (using Pearson correlation coefficient), and 3) Bland-Altman plots to evaluate agreement using both numeric and percentage differences between the methods. We also performed linear regression to explore the relationship between mean ALPS values and differences (both numeric and percentage) in scores between the manual and automatic approaches.

## Results

### Subject characteristics

Our study cohort consisted of 91 patients with PD, 68 patients with iRBD and 50 controls. Out of these, we excluded 12 subjects due to preprocessing failure, including failure of the topup step and the transformation to the normalized space. One patient was excluded due to a structural abnormity. Furthermore, 12 subjects were excluded due to eccentric head position that made the identification of manual ROIs impossible. In total, 25 subjects were excluded. The final yield was 79 subjects with PD, 57 subjects with iRBD, and 48 controls (Table 1).

**Table 1.**

| Table 1 |  |  |  |  |  |  |
| --- | --- | --- | --- | --- | --- | --- |
| Characteristics | Controls (n=48) | iRBD (n=57) | PD (n=79) | Inter-group $p$ value | Partial $\eta^2$ | Between groups |
| Male sex (%) | 34 (71) | 49 (86) | 48 (61) | **0.006** |  | iRBD>>PD |
| Age (years) | 61.5±9.9 | 66.5±7.2 | 59.5±12.0 | **<0.001** |  | iRBD>C; iRBD >>>PD |
| MoCA | 25.6±2.4 | 23.7±2.8 | 24.7±3.1 | **0.031\*** |  | C>iRBD |
| MDS-UPDRS I | 3.0±2.7 | 7.5±5.3 | 5.8±4.3 | **<0.001\*** |  | iRBD>>>C; iRBD>PD;<br>PD>>C |
| MDS-UPDRS II | 0.7±1.5 | 2.6±3.9 | 7.5±4.9 | **<0.001\*** |  | PD>>>iRBD, C;<br>iRBD>C |
| MDS-UPDRS III | 3.4±4.0 | 6.1±5.4 | 30.3±13.1 | **<0.001\*** |  | PD>>>iRBD, C |
| Disease duration** | n.d. | 8.2±8.5 | 2.0±1.8 | n.d. |  | n.d. |
| Manual ALPS scores |  |  |  |  |  |  |
| Average | 1.35±0.17 | 1.31±0.16 | 1.28±0.19 | **0.018\*** | 0.043 | PD<<C |
| Left hemisphere | 1.31±0.16 | 1.27±0.17 | 1.26±0.20 | 0.162* | - | - |
| Right hemisphere | 1.40±0.23 | 1.35±0.18 | 1.31±0.20 | **0.009\*** | 0.051 | PD<<C; PD<iRBD |
| Automatic ALPS scores |  |  |  |  |  |  |
| Average | 1.28±0.11 | 1.24±0.14 | 1.21±0.18 | **0.002\*** | 0.067 | PD<<C, iRBD |
| Left hemisphere | 1.29±0.11 | 1.25±0.14 | 1.22±0.19 | **0.003\*** | 0.063 | PD<<C; PD<iRBD |
| Right hemisphere | 1.27±0.13 | 1.24±0.14 | 1.20±0.18 | **0.003\*** | 0.061 | PD<<C, iRBD |
| **Comparison of demographic and clinical characteristics, and of manual and automatic ALPS scores in PD, iRBD and controls**<br>Where applicable, the data are presented as mean ± standard deviation, with ALPS scores adjusted for age.<br>*MoCA* Montreal Cognitive Assessment, *MDS-UPDRS* Movement Disorder Society-Sponsored Revision of the Unified Parkinson's Disease Rating Scale, *C* controls, > for p<0.05, >> for p<0.01, >>> for p≤0.001, bold for p<0.05<br>*Adjusted for age and sex<br>**Disease duration is defined as subjective duration of dream enactment behavior in iRBD and time since the occurrence of first motor symptom in PD |  |  |  |  |  |  |

The iRBD group was significantly older than both the PD and control group; had a significantly larger portion of males than the PD group; and had significantly lower MoCA scores than the control group. MoCA scores were lower in the iRBD group compared to controls even when controlling for age and sex (p=0.009). The PD group had significantly higher MDS-UPDRS II and III scores than both iRBD and control groups and significantly higher MDS-UPDRS I score than the control group. The iRBD group had significantly higher MDS-UPDRS I and II scores than the control group.

### Reliability analysis of manual and automatic approaches

We compared all three parameters (mean, right-hemisphere, and left-hemisphere) of manual and automatic ALPS score calculation using paired Student’s t-test, bivariate correlation, and drawing a Bland-Altman plot.

Using a paired Student’s t-test, a significant difference was found in the average ALPS scores [t(183)= 9.334, p=<0.001, Cohen’s d=0.41], with manual ALPS scores being significantly higher (mean 1.30, SD 0.18) than the automatic ones (mean 1.23, SD 0.16). This was also true for both the left hemisphere (p=0.001) and right hemisphere (p<0.001) ALPS scores.

Pearson correlation coefficient showed tight association for average [p<0.001, r(182)=0.83], left-hemisphere [p<0.001, r(182)=0.80], and right-hemisphere [p<0.001, r(182)=0.75] ALPS scores. Scatter plots are shown in Supplementary Figure 1.

Bland-Altman plots were drawn using the numeric differences between the manual and automatic approaches, and the linear regression slope was fit (Supplementary Figure 1). Less than 10% of all subjects fell outside of the top and bottom 95% limits of agreement. All three parameters have shown a significant (p<0.05) increase in the difference values with increasing average ALPS values. This increase remained significant when using a percentage difference in the left- and right-hemisphere ALPS score calculation, but not the mean ALPS score.

### Inter-group differences in manual and automatic DTI-ALPS scores

Using one-way ANCOVA on age-adjusted ALPS scores and controlling for sex, we found an inter-group difference in both the manual [F(2,180)=4.081, p=0.018] and the automatic [F(2,180)=6.448, p=0.002] approaches. In the post-hoc analysis, PD subjects had lower ALPS scores using the manual (p=0.008, p=0.051 [PD vs controls, PD vs iRBD]) and automatic (p=0.001, p=0.009 [PD vs controls, PD vs iRBD]) approaches (Figure 2, Table 1).

**Figure 2.**
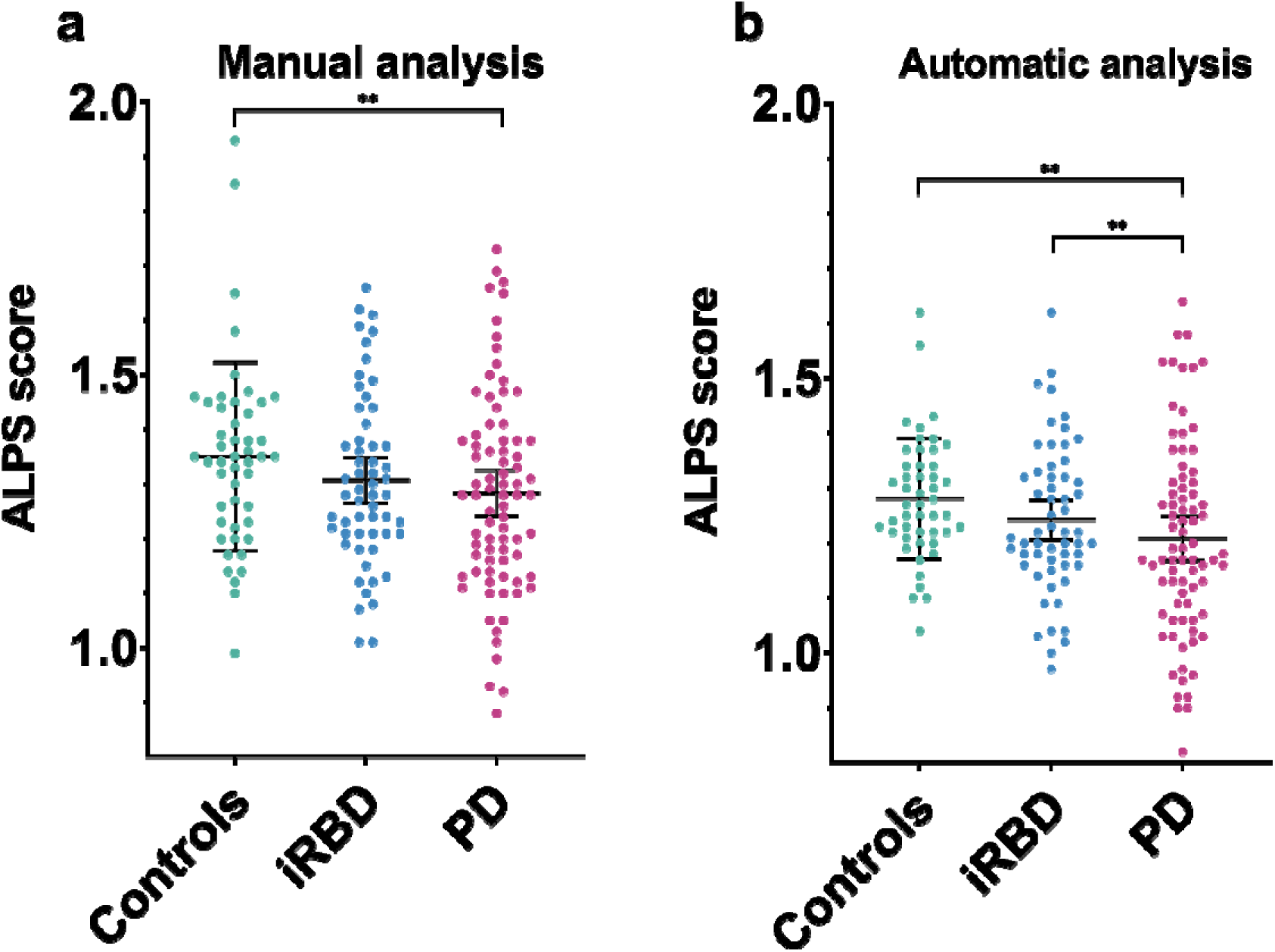
Manual and automatic ALPS score differences between PD, iRBD and HCs. Scatter plot with manual (a) and automatic (b) ALPS scores in PD, iRBD and HCs. Volumes adjusted to age. Line and whiskers represent mean and standard deviations. **<0.01

We also compared the inter-group differences in left- and right-sided age-adjusted ALPS scores, using ANCOVA and controlling for sex. The results were comparable to the averaged values, except for the left-sided manual ALPS scores, where no inter-group difference was found (Table 1).

As the iRBD group was significantly older than the PD group and had significantly more male participants, we performed a sensitivity analysis in age- and sex-matched subgroups. This yielded 40 participants in each group. Using identical methods with age-adjusted average ALPS scores and controlling for sex, we found an inter-group difference in the automatic approach [F(2,116)=5.607, p=0.005], with the PD group having significantly lower ALPS scores (p=0.003, p=0.007 [PD vs controls, PD vs iRBD]). We did not find any significant inter-group differences using the manual approach (p=0.067).

### Effects of nigrostriatal denervation

Using partial correlation on age-adjusted average ALPS scores and controlling for sex, we found a positive correlation between ALPS scores and mean putaminal SBR Z-scores, utilizing both the manual [p=0.029, r(129)=0.191] and the automatic [p=<0.001, r(129)=0.310] approaches in the PD- iRBD pooled group (Figure 3). When analyzing the PD group separately, the results remained significant for both the manual [p=0.027, r(76)=0.250] and the automatic [p=0.011, r(76)=0.286] approaches.

**Figure 3.**
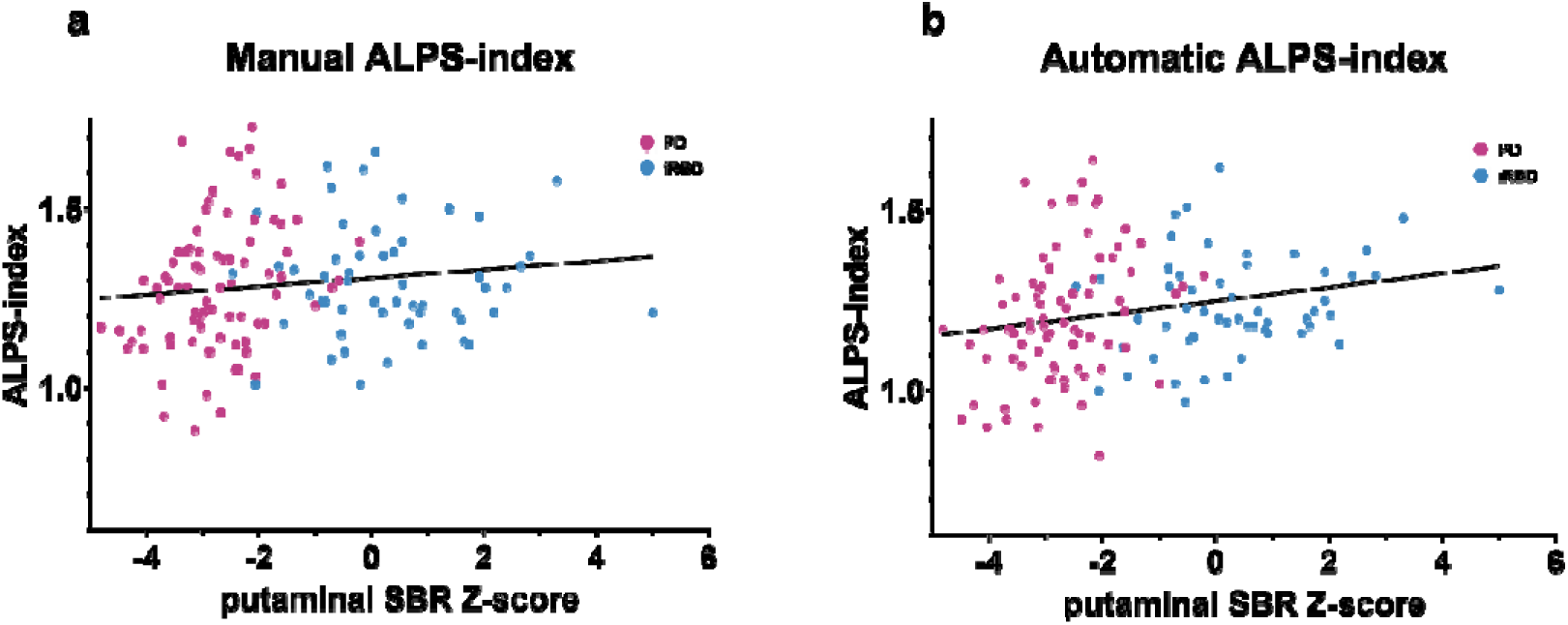
Manual and automatic ALPS score in relation to putaminal SBR Z-scores. Scatter plots representing the relation of manual (a) and automatic (b) ALPS scores with putaminal SBR Z-scores. Volume to age. PD and iRBD subjects are pooled.

We further compared ALPS scores in the most affected and least affected hemisphere with regard to putaminal SBR on DAT-SPECT. We did not find any significant differences between the most and least affected hemisphere in the pooled group or when PD and iRBD groups were analyzed separately.

### Associations with symptoms severity

Using partial correlation on age-adjusted ALPS scores and controlling for sex, we found negative correlations of ALPS scores with several MDS-UPDRS scores in PD (Table 2). The manual approach correlated with MDS-UPDRS I score. Both manual and automatic methods correlated with MDS-UPDRS II and III scores, and ALPS scores also correlated with the bradykinesia and axial MDS-UPDRS III subscores in PD.

**Table 2.**

| Table 2 |  |  |  |  |  |  |  |
| --- | --- | --- | --- | --- | --- | --- | --- |
|  | MDS-UPDRS I | MDS-UPDRS II | MDS-UPDRS III | Tremor | Rigidity | Bradykinesia | Axial |
| Manual | **0.005(-0.32)** | **<0.001(-0.45)** | **0.002(-0.35)** | 0.665(-0.05) | 0.135(-0.172) | **<0.001(-0.41)** | **0.003(-0.34)** |
| Automatic | 0.055(-0.22) | **0.001(-0.36)** | **0.028(-0.25)** | 0.626(0.06) | 0.220(-0.14) | **0.006(-0.31)** | **0.001(-0.37)** |
| **Associations of manual and automatic ALPS scores with symptom severity in PD**<br>Using two-tailed partial correlation on age adjusted ALPS scores, controlling for age and sex.<br>Values presented as p-value(partial correlation coefficient).<br>Bold for p<0.05 |  |  |  |  |  |  |  |

Using sex as a covariate, we did not find any significant correlation between age-adjusted ALPS scores and MoCA scores in any of the groups.

## Discussion

Our results show a lower ALPS-index, representing decreased diffusion in the direction of glymphatic flow, in subjects with PD, compared to subjects with iRBD and healthy controls. Moreover, this decrease correlates with nigrostriatal denervation, as measured by DAT-SPECT, and clinical severity, as measured by MDS-UPDRS clinical scales.

DTI-ALPS, first described by Taoka et al. in 2017^13^, is a relatively new approach to measure glymphatic system dysfunction. As such, the method still lacks rigorous pathological verification^6,34^ proving that ALPS-index represents the actual magnitude of glymphatic flow.

One of the other critiques of the DTI-ALPS method is the possibility of human error during ROI selection process. We have addressed this issue by incorporating an automatic ROI selection approach, which, based on our findings, demonstrated performance comparable to or superior to the manual method. Our approach was similar to other studies that employed atlas method of ROI selection, with one study also using a similar JHU atlas^35^ and others using an ICBM DTI-81 atlas^36,37^.

We chose to employ the JHU atlas, as it was readily available in the FSL installation used in the preprocessing steps.

When comparing the manual and automatic DTI-ALPS calculation methods, we found that manual ALPS scores were significantly higher than the automatic ones. When further analyzed using the Bland-Altman plots, we found an increasing difference between the methods with increasing ALPS score values, with only a small number of subjects falling outside the upper and lower 95% limits of agreement in all Bland-Altman plots. However, the correlation between the methods was excellent.

In our study, all DTI-ALPS scores (mean, left, right, manual, and automatic) showed similar results, except the left-hemisphere manual ALPS scores, which did not show significant between-group differences. This finding is unexpected, as the placement of left-sided association ROIs is generally easier, since the SLF of right-handed subjects is better discernible on the usually dominant left side (as most of the population is right-handed)^6^. This could be due to human error during ROI selection, which highlights the need for automated ROI selection. Future studies should verify the performance of different atlases in automatic DTI-ALPS calculation.

We found two previous studies comparing PD, iRBD, and healthy control subjects. Both found significant differences in the ALPS-index between iRBD and control subjects. One study included “probable” iRBD (cases not confirmed by polysomnography) and found significant differences between all three groups^14^. The other one included iRBD subjects confirmed by polysomnography and found significant differences between iRBD and HC, not iRBD and PD subjects, however it included only 20 patients in each group^9^. Some previous studies compared ALPS scores in iRBD and control subjects only and found significantly reduced ALPS scores in iRBD subjects^8^. In our study, we have not confirmed this finding. This could be due to several reasons, including inter-rater variability in the selection of manual ROIs used in these studies or differences in the patient populations and relatively low number of iRBD patients leading to limited statistical power. Our iRBD subjects were recruited “on-demand” by a media campaign. This difference of patients being approached versus patients spontaneously seeking medical help for the symptoms of their iRBD could, in our case, select for a different, perhaps earlier stage, patient population than in previous studies. However, the difference in ALPS scores between iRBD and PD was expected, as PD is a later stage of synucleinopathy and neurodegeneration than iRBD.

We found two studies comparing ALPS scores to DAT-SPECT. One study involved drug-naïve PD patients, while another focused on individuals with iRBD, neither finding a correlation between ALPS scores and DAT-SPECT.^9,38^ Another study found a correlation between nigral dopaminergic degeneration and ALPS-scores using a hybrid 18F-fluorodopa PET-MRI^39^. Our results show that even using a more simple approach to nigrostriatal denervation imaging, ALPS scores were correlated with the degree of nigrostriatal denervation in PD but not iRBD subjects. This implies that glymphatic system dysfunction may be a progression marker, similar to nigrostriatal denervation, both being dynamic processes worsening with the disease stage. As no correlation was found in the iRBD group, the glymphatic dysfunction could be a more late-stage pathological process than the nigrostriatal denervation; or the changes could be less pronounced in the earliest stages of neurodegeneration. The lack of a significant PD-iRBD between-group difference in ALPS scores in our study supports this. Future studies will be needed to assess the extent to which glymphatic system dysfunction is accelerated by synucleinopathy, compounding the “physiological” age-related decline in glymphatic flow.

An interesting finding is the correlation of ALPS index with MDS-UPDRS scores and bradykinesia and axial MDS-UPDRS III subscores. The correlation with MDS-UPDRS scores has been described before^38,40^. In a previous study, the results differed, with ALPS scores correlating with only the rigidity subscores^41^.

Our study did not find a correlation between ALPS scores and MoCA. There are inconsistent results regarding the correlation between cognitive dysfunction and ALPS scores. Some studies did not find a correlation between ALPS scores and MoCA^14,41^, with others describing a relationship between cognitive dysfunction and ALPS scores^5^. However, our subjects were in a very early stage of the disease. As overt cognitive deficit is not a feature of iRBD or generally of early-stage PD, it is possible that the association would appear at later stages, where a larger spread of MoCA scores would be available due to an onset of dementia in a subset of patients.

We also did not find a side difference between the side more and the side less affected by the nigrostriatal denervation. This can be explained by the glymphatic dysfunction being a global process affected by overall neurodegeneration, not “local” side differences in nigrostriatal denervation.

In conclusion, our results support a body of evidence pointing to glymphatic dysfunction in PD. The glymphatic dysfunction correlates with clinical severity as measured by MDS-UPDRS scores and nigrostriatal denervation as measured by DAT-SPECT. This suggests that glymphatic dysfunction is a dynamic process that worsens with disease progression. The ALPS scores of iRBD patients were numerically intermediate between those of the PD and HC groups, with significant differences observed only between iRBD and PD patients. Moreover, we have shown that an automatic approach to ALPS calculation is comparable, and in some cases superior, to the manual approach.

## Data Availability

The datasets used and analyzed during the current study are available from the corresponding author on request.

## Acknowledgements

### Study funding

The study was supported by: National Institute for Neurological Research (Programme EXCELES, ID Project No. LX22NPO5107) - Funded by the European Union – Next Generation EU; General University Hospital in Prague project MH CZ-DRO-VFN64165 and Na Homolce Hospital project CZ- DRO-NHH00023884 and Czech Health Research Council grant NU21-04-00535. Computational resources were provided by the e-INFRA CZ project (ID:90254), supported by the Ministry of Education, Youth and Sports of the Czech Republic. Access to CESNET storage facilities provided by the project „e-INFRA CZ“ under the programme „Projects of Large Research, Development, and Innovations Infrastructures“ LM2018140), is greatly appreciated.

### Authors’ Roles

S.M. co-designed the study concept, implemented the se preprocessing and automatic segmentation algorithm, performed statistical analysis, and wrote the draft of the manuscript. V.R. co-designed the study concept and managed the manual placement of ROIs. J.N. co-designed the study concept and performed the polysomnography analysis. T.K. managed the resource-intensive computing on MetaCentrum and automated the algorithm. R.K. advised on automated algorithm creation and statistical analysis. J.K. contributed to MRI acquisition. D.Z. contributed to DAT-SPECT acquisition and processing. J.T. contributed to DAT-SPECT acquisition and processing. K.S. contributed to the polysomnography acquisition and processing, and to the critical edition of the manuscript. E.R. contributed to the clinical data acquisition and to the critical edition of the manuscript. P.D. co- designed the study concept, performed expert supervision, clinical examination, contributed to MRI acquisition, and to the critical edition of the manuscript.

### Financial disclosure and Conflict of Interest

The authors declare that there are no financial disclosures or conflicts of interest relevant to this work.

## Notes

### Competing Interest Statement

The authors have declared no competing interest.

### Author Declarations

Ethics committee of the General University Hospital in Prague gave ethical approval for this work.

