## Supplementary Figure 1 for "From iRBD to Parkinson’s disease: Tracking Glymphatic Dysfunction Using Automated DTI-ALPS Analysis"

**Analysis of differences in manual and automatic ALPS score calculations**

Scatter plots representing the relation of manual and automatic DTI-ALPS calculation approaches (a-c). Bland-Altman plots using the numeric difference values (d-f) and % difference values (g-I) between the manual and automatic approaches.

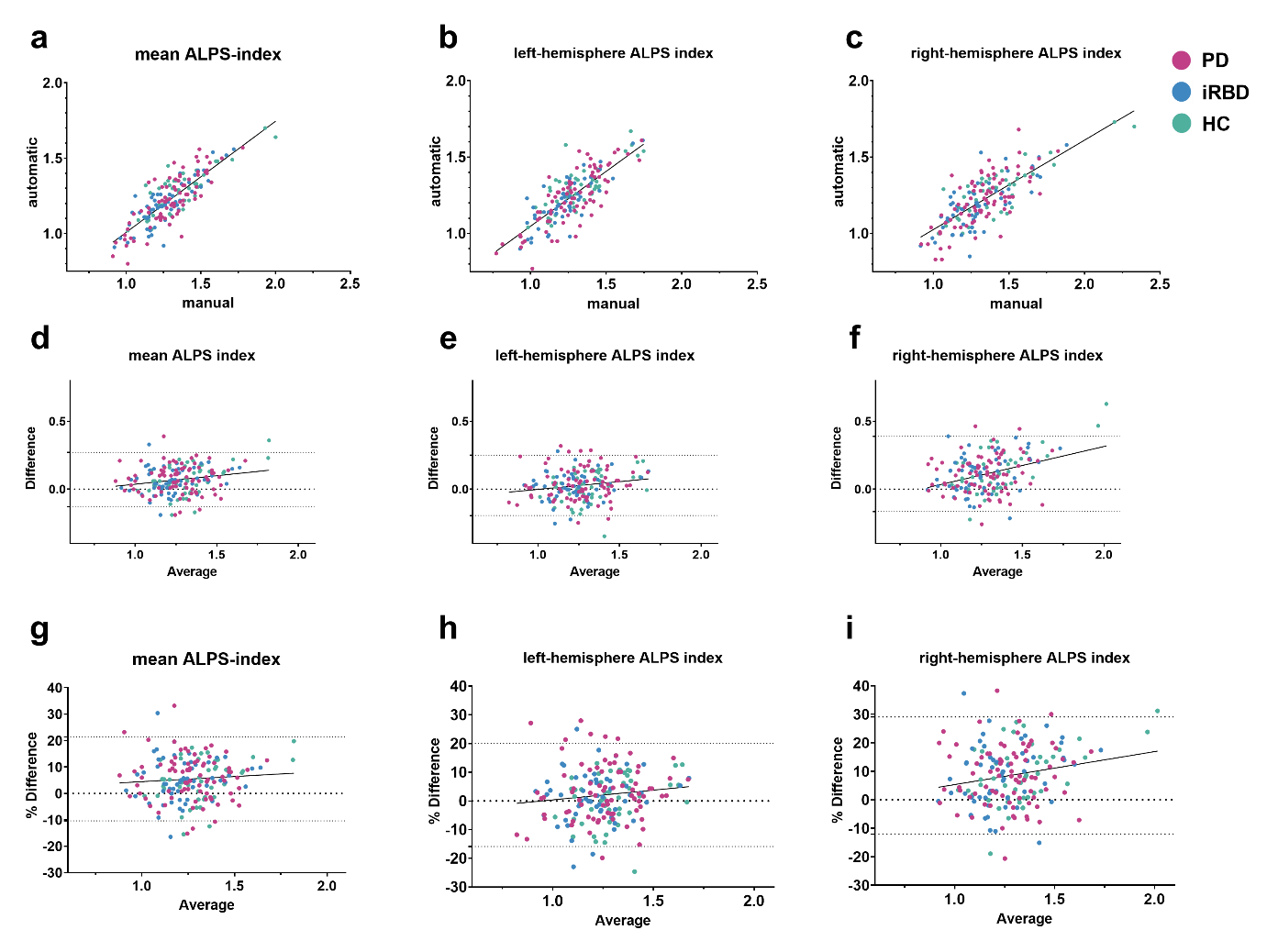
